# Plasma neurofilament light chain is elevated in Niemann-Pick Type C but glial fibrillary acidic protein is not

**DOI:** 10.1101/2023.12.12.23299884

**Authors:** Dhamidhu Eratne, Courtney Lewis, Wendy Kelso, Samantha Loi, Wei-Hsuan Michelle Chiu, Kaj Blennow, Henrik Zetterberg, Alexander F Santillo, Dennis Velakoulis, Mark Walterfang, And The MiND Study Group

## Abstract

**OBJECTIVE:** Niemann-Pick Type C (NPC) is a genetic neurodegenerative lysosomal storage disorder commonly associated with psychiatric symptoms and delays to accurate diagnosis and treatment. This study investigated biomarker levels and diagnostic utility of plasma neurofilament light chain (NfL) and glial fibrillary acidic protein (GFAP) in NPC compared to healthy controls.

**METHODS:** Patients with NPC were recruited from a specialist assessment and management service. Data was available from an age and sex-matched healthy control group. NfL and GFAP were measured on Quanterix Simoa HD-X analysers and groups compared using genralised linear models. NfL levels were compared to, and percentiles derived from, recently developed NfL reference ranges.

**RESULTS:** Plasma NfL was significantly elevated in 11 patients with NPC compared to 25 controls (mean 17.1pg/mL vs 7.4pg/mL, p<0.001), and reference ranges (all >98^th^ percentile). NfL distinguished NPC from controls with high accuracy. GFAP levels were not elevated in NPC (66.6pg/mL vs 75.1pg/mL).

**DISCUSSION:** The study adds important evidence on the potential diagnostic utility of plasma NfL in NPC, extends the literature of NfL as a diagnostic tool to differentiate neurodegenerative from primary psychiatric disorders, and adds support to the pathology in NPC primarily involving neuronal, particularly axonal, degeneration.

## INTRODUCTION

Niemann-Pick Type C (NPC) is a rare, severe, genetic neurodegenerative lysosomal storage disorder, associated with highly variable age at onset and symptoms. NPC is commonly associated with psychiatric symptoms, high rates of misdiagnosis and delay until accurate diagnosis and treatment.^1–3^ While there are no specific blood or cerebrospinal fluid (CSF) biomarkers which assist in the diagnosis of NPC, biomarkers of neuronal injury (e.g., neurofilament light chain protein, NfL), and astrocytosis (e.g., glial fibrillary acidic protein, GFAP) may be useful as non-specific markers. Elevated NfL and GFAP levels have been identified in neurodegenerative conditions such as Alzheimer disease and frontotemporal dementia,^4–6^ and NfL has been shown to distinguish neurodegenerative from primary psychiatric and non-neurodegenerative conditions, and potentially reduce misdiagnosis.^6–10^

Studies have found elevated CSF and plasma NfL levels in NPC compared to primary psychiatric conditions and controls,^11,12^ and associations with severity and miglustat treatment.^13^ No studies have investigated GFAP in NPC, except one that included only two people with NPC as a comparison group, not finding elevated levels.^14^

This study aimed to compare NfL and GFAP levels in patients with NPC to age- and sex-matched controls, and to compare NfL levels to the reference ranges we recently developed. ^8^ Exploratory analyses investigated associations with clinical variables and explored biomarker levels in patients who had serial bloods.

## METHODS

Patients were recruited from a tertiary specialist NPC assessment and management service at Neuropsychiatry Centre, Royal Melbourne Hospital, Australia. Data was available from an age-matched healthy control group with no cognitive, neurological, or psychiatric symptoms or conditions, no known renal impairment or other severe medical conditions. Plasma aliquots were collected, processed, and stored at -80°C. Plasma NfL and GFAP levels were measured on Quanterix Simoa HD-X analysers.

Statistical analyses were performed using R v4.2.2 (2022-10-31). Generalised linear models (GLMs) were used to examine relationships with log10-transformed biomarker levels, diagnostic group, age and sex as covariates, with 95% confidence intervals (nonparametric bootstrapping, 1000 replicates). Receiver operator characteristic curves were computed to estimate diagnostic performance, area under the curve (AUC), sensitivity, and specificity. For more precise exploration of NfL levels, levels were compared to and percentiles derived from reference ranges developed using generalised additive models for location, scale, and shape, previously described.^8^

This study, part of The Markers in Neuropsychiatric Disorders Study (The MiND Study, https://themindstudy.org), was approved by the Melbourne Health Human Research Ethics Committee (MH/HREC2020.142).

## RESULTS

There were 11 patients with NPC, all of whom had neurological symptoms, and 25 controls. There were no differences in age (mean age 33.9 years vs. 40.7, p=0.098) or sex (63.6% vs. 80%, p=0.409). The mean duration of illness was 12.2 years. Four out of 11 (36%) patients had a juvenile onset; 7/11 (64%) were on treatment (details in Table 1).

**Table 1.** Study demographics and plasma biomarker levels.

|  | Control<br>*N*=25 | NPC<br>*N*=11 | p.overall |
| --- | --- | --- | --- |
| Age at sample, years | 40.7 (7.44) | 33.9 (11.8) | 0.098 |
| Sex, females | 20 (80.0%) | 7 (63.6%) | 0.409 |
| Age at onset, years | - | 20.8 (9.58) |  |
| Duration of illness, years | - | 12.2 (5.52) |  |
| Juvenile or adult onset: |  |  |  |
| Adult | - | 7 (63.6%) |  |
| Juvenile | - | 4 (36.3%) |  |
| NfL, pg/mL | 7.44 (2.62) | 17.1 (5.90) | <0.001 |
| LogNfL | 0.85 (0.13) | 1.21 (0.15) | <0.001 |
| GFAP | 75.1 (27.6) | 66.6 (16.6) | 0.262 |
| LogGFAP | 1.85 (0.17) | 1.81 (0.10) | 0.460 |
| On Treatment: |  |  |  |
| No | - | 4 (36.4%) |  |
| Yes | - | 7 (63.6%) |  |
| Treatment detail: |  |  |  |
| Miglustat | - | 2 (18.2%) |  |
| Miglustat and IV cyclodextrin | - | 2 (18.2%) |  |
| Miglustat and acetylleucine | - | 2 (18.2%) |  |
| Nil treatment | - | 4 (36.4%) |  |
| Acetylleucine | - | 1 (9.09%) |  |

Data is mean (SD) or n (%). GFAP: glial fibrillary acidic protein; NfL: neurofilament light chain protein

### Plasma NfL and GFAP in Niemann-Pick Type C compared to controls

NfL levels were significantly higher in patients with NPC compared to controls (mean 17.1pg/mL vs. 7.4pg/mL, GLM for logNfL: β=1.81, 95% confidence interval: [1.50, 2.22], p<0.001), Table 1 and Figure 1. As demonstrated in Figure 2, 100% of patients with Niemann-Pick Type C had significantly elevated NfL levels for their age: all were >98^th^ percentile (z-score>2); 7/11 (64%) were >99^th^ percentile (z-score>2.5).

**Figure 1.**
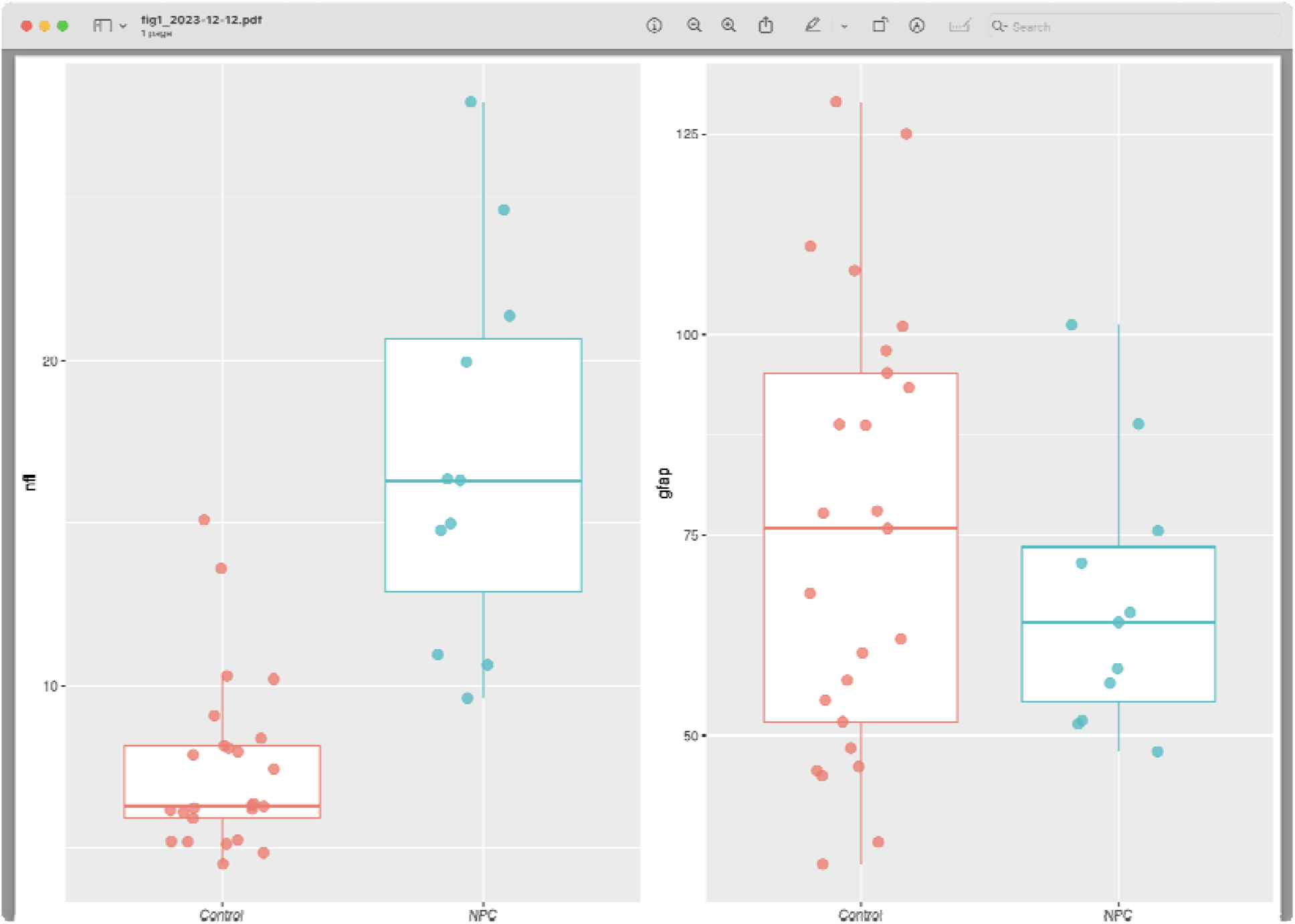
**Plasma neurofilament light chain protein (NfL) and glial fibrillary acidic protein (GFAP), levels in pg/mL, in patients with Niemann-Pick Type C (NPC) compared to controls.** High quality file for upload: fig1_2023-12-12.pdf

**Figure 2.**
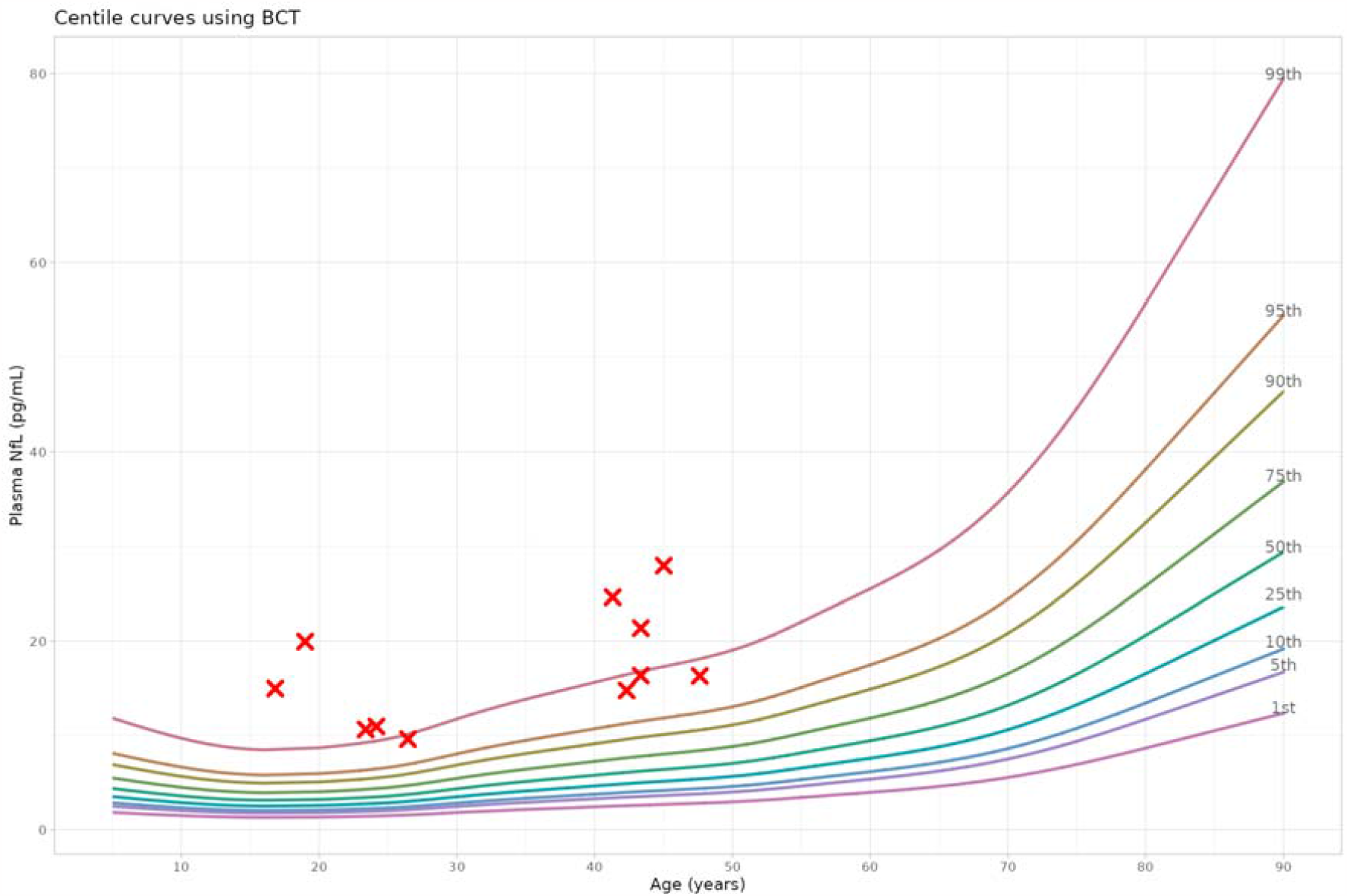
**All patients with Niemann-Pick Type C had significantly elevated NfL levels for their age (all >98-99^th^ percentile), using our previously developed interactive plasma NfL reference range app (themindstudy.org/apps)** High quality file for upload: fig3_2023-11-21.png

By comparison, GFAP levels were not elevated in NPC compared to controls (66.6pg/mL vs. 75.1pg/mL, β=-0.09 [-0.83, 0.63], p=0.790).

Plasma NfL had high diagnostic accuracy to distinguish NPC from controls (AUC 0.96 [0.91, 1.00]). An optimal cut-off of 9.35pg/mL was associated with 84% specificity, 100% sensitivity. A cut-off of 10.47pg/mL resulted in better specificity (92%), but slightly reduced sensitivity (91%). GFAP did not have diagnostic utility (AUC 0.57 [0.38, 0.76]).

### Exploratory analyses

Exploratory analyses investigated biomarker levels differences between treated versus untreated patients, different types of treatment, and juvenile versus adult onset. No differences were seen.

Three patients had serial bloods. Over a two-year period, NfL levels in patient A (on miglustat and IV cyclodextrin) increased only slightly, by 11% (from 9.6pg/mL to 10.7pg/mL), while GFAP increased by 36% (from 88.8pg/mL to 121pg/mL). On the other hand, patient B (only on miglustat and acetylleucine), exhibited a 50% increase in NfL over 2 years (10.6pg/mL to 16pg/mL), and a 76% increase in GFAP (65.3pg/mL to 114.8pg/mL). Patient C (on acetylleucine only), had a significant neurological deterioration requiring hospitalisation at the time of their second blood sample. Their second level (58pg/mL) was markedly higher than a year prior (20pg/mL), and GFAP increased by 28% (51.4pg/mL to 66pg/mL).

## DISCUSSION

This study found significantly elevated plasma NfL levels in NPC compared to controls, and NfL distinguished NPC from controls with high accuracy. These findings add important evidence on the potential diagnostic utility of plasma NfL in this devastating condition that is not uncommonly misdiagnosed as primary psychiatric conditions. In addition, this study provides an important negative finding of plasma GFAP in the largest number of NPC patients to date.

The differing profile of NfL and GFAP adds weight to the pathology in NPC primarily involving neuronal, and particularly axonal, degeneration. ^15^ It is likely that varying profiles of these biomarkers will be seen in different disorders, providing important information on the underlying pathophysiological processes in various conditions and within subgroups of syndromes, and combination biomarkers could have diagnostic and wider clinical utility in various conditions and differential diagnoses.

Secondary, purely exploratory analyses did not find any differences in NfL and GFAP levels between untreated and treated patients, however the main limitation of this study is the small sample size and lack of serial levels, which limit any confident interpretations of treatment effects and changes over time. Three patients with serial levels had large increases in GFAP levels over 1-2 years. One of these two patients was on IV cyclodextrin and did not have much increase in NfL, whereas the other had a more marked increase. This may reflect a treatment effect of IV cyclodextrin on neuronal injury, however, this was not seen in a larger study. ^13^ The increase in GFAP over time is notable, possibly pointing to differing timelines of NfL and GFAP changes in the disease course, with NfL changing earlier, and GFAP changing later, perhaps after neuronal loss has occurred. Patient C’s dramatic increase in serial NfL levels corresponded with severe neurological deterioration. This adds weight to NfL reflecting severe/acute deterioration and severity/prognosis/rate of deterioration, on top of being diagnostic. Our cohort included patients with several years of symptoms, therefore we cannot conclude the diagnostic utility at the earliest stages of the disease. It would be important to investigate serial biomarkers and associations with other variables in larger, well characterised cohorts.

To conclude, this study found significantly higher plasma NfL levels in NPC and demonstrated strong diagnostic utility of plasma NfL to distinguish NPC from controls, while not finding elevated GFAP levels. This extends the literature on NfL to identify neurological/neurodegenerative causes of neurological and neuropsychiatric symptoms, especially in younger people where diagnostic challenges and misdiagnosis can be higher, and adds to our understanding of the pathophysiology and utility of biomarkers in NPC for diagnosis and potentially wider clinical utility. Further studies are underway in a larger cohort, to investigate NfL, GFAP, and other biomarkers and associations with clinical and treatment variables.

## Data Availability

All data produced in the present study are available upon reasonable request to the authors

